# Social network-based strategies for classroom size reduction can help limit outbreaks of SARS-CoV-2 in high schools. A simulation study in classrooms of four European countries

**DOI:** 10.1101/2020.11.30.20241166

**Authors:** Anna Kaiser, David Kretschmer, Lars Leszczensky

## Abstract

Dividing classrooms may reduce the risk of SARS-CoV-2 outbreaks in schools. We investigate how classroom cohorting strategies, which downsize and isolate groups, may curb the spread of SARS-CoV-2. Using agent-based modelling based on a rich multi-country network dataset comprising 507 classrooms and 12,291 students, we assess random cohorting and three network-based strategies that consider students’ out-of-school contacts with classmates. Investigating effects on the number of cross-cohort transmissions, overall infections, and quarantines, our findings suggest that all cohorting strategies help to contain outbreaks, but that minimizing out-of-school contact between cohorts is most effective. Since this strategy may be hard to implement in practice, we show that a network chain nomination procedure and splitting classes by gender, both of which are easier to realize, also outperform random cohorting considerably. For all cohorting strategies, we find that rota-systems with instruction in alternating weeks contain outbreaks more effectively than same-day in-person instruction.

---

Schools have long been identified as drivers of influenza and other respiratory-spread epidemics^1–5^. When the novel coronavirus SARS-CoV-2 resulted in a pandemic in early 2020, many countries reacted by closing schools. However, school closures are controversial both in terms of their effect on disease transmission and because of their social and economic costs.

Modeling studies indicate that school closures may mitigate the spread of SARS-CoV-2, but the exact degree to which school closures reduce transmission is not known^6–16^. School closures were often synchronized with other non-pharmaceutical interventions, such that measuring their individual effect is challenging. Two recent multi-country studies that disentangle the effects of different non-pharmaceutical interventions find school closures to stand out as one of the most influential measures^17,18^. Observational data suggests that school-based outbreaks have been infrequent in the summer and early fall of 2020^19–24^, but incidence was low in most countries in this time period and larger infection clusters have also been reported^25–27^. An epidemiological assessment of the role of schools in the SARS-CoV-2 pandemic is further complicated by uncertainty surrounding children’s propensity to transmit the disease. Emerging evidence on a lower force of infection in children under the age of ten suggests that primary schools and childcare facilities may be at lower risk. However, secondary schools may be at higher risk because transmission appears to be stronger in adolescents, possibly approximating transmission dynamics in adults^28–32^.

Though potentially reducing SARS-CoV-2 transmission, school closures have adverse social and economic consequences. In the short term, they raise the burden on working parents, including those in health-care professions who are indispensable in a pandemic^33,34^. In the long run, school closures are associated with negative learning outcomes^35,36^, as distance learning seems to be an insufficient substitute for in-person classes^37,38^, especially for those from socio-economically disadvantaged backgrounds^35^. For these reasons, school closures are seen as a last resort in public debates.

To delay or avoid school closures, strategies that reduce the probability of infections and the size of infection clusters in schools are necessary. Schools mainly facilitate the spread of communicable diseases by bringing together large numbers of interconnected individuals. Therefore, social distancing strategies that decompose the student population into smaller isolated units may be effective to reduce the risk of large infection clusters. However, apart from one modelling study on group size reductions in US high schools^39^, there is a surprising lack of research on social distancing measures during SARS-CoV-2 outbreaks in schools^32^. In the European context that we focus on, in-school instruction is typically organized in classrooms of 20-40 students in which most courses are taught to the same set of students. Under these conditions, we examine social distancing strategies that divide these classrooms into smaller isolated units that are instructed separately, aiming to avoid large clusters and super-spreading events.

In line with the definition of the Centre for Disease Control and Prevention^40^, we term the process of splitting classes *cohorting* and the resulting separate groups *cohorts*. We consider cohorting strategies that split full classrooms (i.e., about 20-40 students) into two cohorts of approximately equal size. Cohorting classrooms has several benefits for preventing the transmission of SARS-CoV-2. It facilitates physical distancing within the classroom because there is more space per student and it reduces the number of students who are exposed to an infection within the classroom, which can moderate the size and the reach of an initial outbreak.

Reorganizing teaching and dividing classrooms is costly. Therefore, it is important to rely on cohorting strategies that prevent the transmission of SARS-CoV-2 between cohorts as effectively as possible. However, preventing transmission between cohorts requires to not only isolate cohorts *within* the school context, but also *outside* of school. Therefore, the effectiveness of cohorting can likely be improved by accounting for out-of-school contact networks among students. We compare the effect of such network-based cohorting strategies to strategies that do not consider out-of-school contact. Our network-based strategies exploit the fact that real-world social networks mostly consist of clusters that are well-connected internally and are more loosely connected with other clusters^41^. This pattern results from the ways in which people form relationships. For example, people tend to be segregated in terms of many characteristics, frequently associating with others who are similar to them^42^. Structural mechanisms are also at work, such as when friends of friends tend to become friends^43,44^. When social networks are organized in clusters with few ties between them, social distancing measures that successfully sever between-group ties should be more efficient than strategies that do not take social network structure into account^45–48^. Our network-based cohorting strategies apply this insight to the spread of SARS-CoV-2 in the school context.

We compare four cohorting strategies to a baseline scenario where classrooms are not divided into cohorts (see table 1 for a summary). The first strategy is ***random cohorting***. In this strategy, classrooms are randomly divided into two equally-sized cohorts. Random cohorting—like all other cohorting strategies—prevents interaction with members of other cohorts within school. However, it does not account for students’ social networks, so that out-of-school contacts that span cohorts can still serve as transmission channels between cohorts.

**Table 1:** Overview of cohorting strategies and key epidemiological outcome measures

| *Cohorting strategies* |  |
| --- | --- |
| Strategy | Description |
| Random cohorting | Two cohorts are formed by randomly allocating half of the students to each cohort. |
| Gender-split cohorting | One cohort consists of boys, one of girls. Students from the smaller cohort (i.e., the underrepresented gender) are reallocated until both cohorts have the same size. (See Supplementary Material for variations.) |
| Optimized cohorting | Two equally-sized cohorts are formed to minimize the number of cross-cohort out-of-school contacts. |
| Network chain cohorting | An initial student names all of her out-of-school contacts, who themselves name their out-of-school contacts, etc., until the resulting set of students comprises half of the classroom. This set of students forms the first cohort, the remainder the second cohort. |
| *Key epidemiological outcome measures* |  |
| Proportion of simulations with transmission to second cohort | Does SARS-CoV-2 spread from the seed node's cohort to the other cohort, such that containment fails? |
| Average proportion of students in quarantine at the end of the simulation | How many students are quarantined and can thus (temporarily) not attend school? |
| Average proportion of students infected at the end of the simulation | How many students in the classroom (across cohorts) have been infected (by the seed node or other students)? |

Our first network-based strategy splits cohorts by gender, exploiting strong gender segregation in adolescents’ networks^49,50^, so that many resulting out-of-school contacts are within rather than between cohorts. This ***gender-split cohorting*** strategy is easy to implement, but cross-gender friendships and an elevated transmission risk in cross-gender romantic relationships may sometimes undermine its efficiency. The second strategy is network-based ***optimized cohorting***. This strategy explicitly uses information on students’ out-of-school contacts with classmates to form cohorts in a way that minimizes the number of cross-cohort contacts. By definition, this strategy produces the cleanest separation of cohorts and should thus be most effective in preventing cross-cohort infection. However, this strategy is hard to implement, as teachers need to know students’ out-of-school contact networks and optimize cohorts accordingly. We therefore propose a ***network chain cohorting*** strategy that uses an in-class nomination procedure to approximate the optimization strategy and is much easier to implement. In this strategy, an initial student who is well-connected—such as a class representative or a student known to be popular—names all of her in-class out-of-school contacts, and the resulting set of students forms the basis for the first cohort. Subsequently, the listed out-of-school contacts name *their* out-of-school contacts, who also become members of the first cohort. The process continues until half of the classroom is allocated to the first cohort, and the remaining students form the second cohort. If the chain breaks, another random student can be allocated to the group and selected to nominate her out-of-school contacts.

Using an exemplary classroom in the dataset, Figure 1 demonstrates both how these four different cohorting strategies induce different cohort compositions and how they can help avoiding cross-cohort out-of-school contacts. While there are many cross-cohort contacts under random cohorting, gender-split and network chain cohorting produce fewer cross-cohort ties and optimized cohorting succeeds in perfectly separating cohorts in this example classroom.

**Figure 1:**
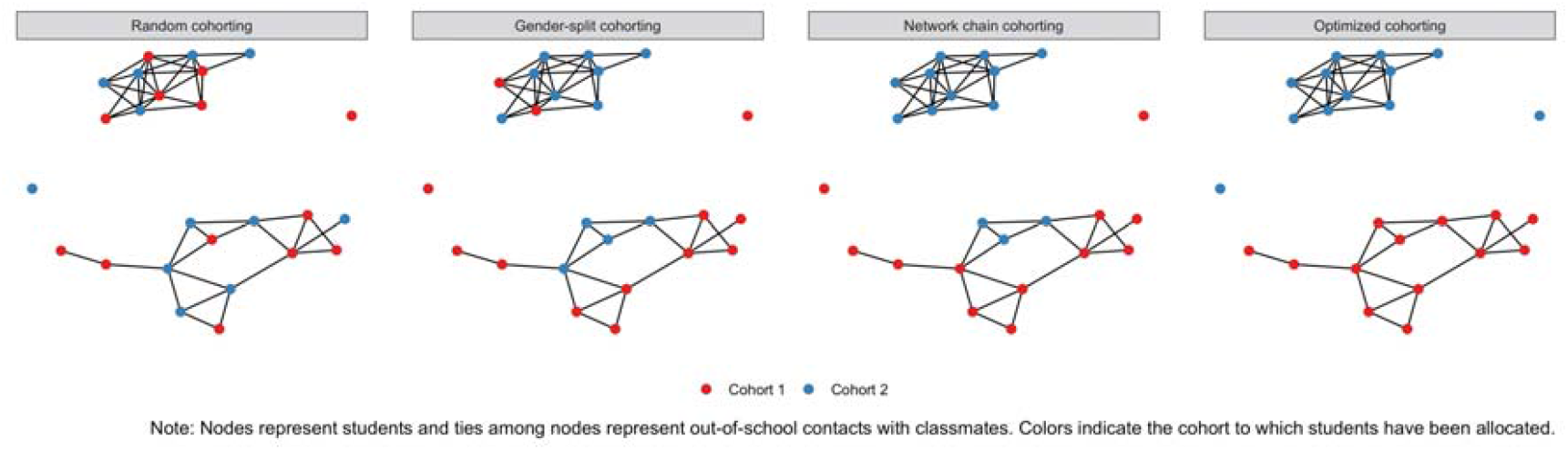
Cross-cohort out-of-school ties for different cohorting strategies in an example classroom from the CILS4EU data.

Apart from the necessity to choose a specific cohorting strategy, cohorting also has consequences for the organization of in-person teaching. In principle, all cohorts can be instructed on the same day, using multiple classrooms or different schedules. Alternatively, cohorts can be taught in a rota-system, with each cohort in turn being instructed in school and online on alternating days, in alternating weeks or in episodes of two weeks. A rota-system reduces the total amount of in-person instruction, but may act as a “natural quarantine” because it allows transmission within cohorts only in certain time intervals. We model a rota-system with in-person instruction of cohorts every other week because it combines “natural quarantines” with the pedagogical benefits of steady in-school face-to-face instruction. In our analysis, we therefore compare the epidemiological consequences of this rota-system to in-person teaching on the same day.

We use simulation models to investigate whether cohorting strategies can help to prevent the transmission of SARS-CoV-2 in classrooms. We compare the effectiveness of the different cohorting strategies in terms of three key indicators that are also summarized in Table 1. First, we consider the proportion of simulations in which SARS-CoV-2 is transmitted to the second cohort, meaning that containment of an initial outbreak within one cohort fails due to transmission via out-of-school contacts. Second, we assess the proportion of students who are quarantined and thus temporarily cannot participate in school activities in person. Third, we evaluate the overall proportion of students in the classroom who become infected to assess whether cohorting strategies can help to reduce the severity of outbreaks.

To simulate the transmission of SARS-CoV-2 in classrooms, we use real-world student network data from the first wave of the Children of Immigrants Longitudinal Study in Four European Countries (CILS4EU) project^51,52^. The data were collected in 2010-2011 and provide information on 14-15-year-old students from England, Germany, the Netherlands, and Sweden. In total, our sample consists of 507 classrooms populated by 12,291 students (for details see methods). Out-of-school interaction is captured by an indicator assessing the classmates a student “often spend[s] time with outside school”. Students could nominate as many of their classmates as they wanted. Whenever one student named another, we code an out-of-school contact between this pair of students, independent of whether the second student confirmed the nomination because contact necessarily goes both ways. The median number of out-of-school contacts is three and the average is 3.58 classmates. In contrast to many other social-network-based transmission models, our study uses complete real-world network data. This allows for representing network structures according to how they are empirically observed among students, including the clustering patterns that make tailored social distancing strategies particularly effective.

In line with previous applications^6,13,39,45,46,53^, we use agent-based modeling to simulate disease trajectories and the transmission of SARS-CoV-2 through in-class interaction and out-of-school contact, summarized in Figure 2 (details on all model steps can be found in the methods section). Analogous to classical epidemiological models, students in our simulations may transition from being susceptible to being exposed, infectious and, eventually, recovered. We simulate transmission dynamics separately for each classroom and each cohorting strategy, repeating each simulation 2000 times due to its stochastic nature. The simulation ends when all students have been infected or quarantined, or when seven weeks have passed (capturing the effect of school holidays).

**Figure 2:**
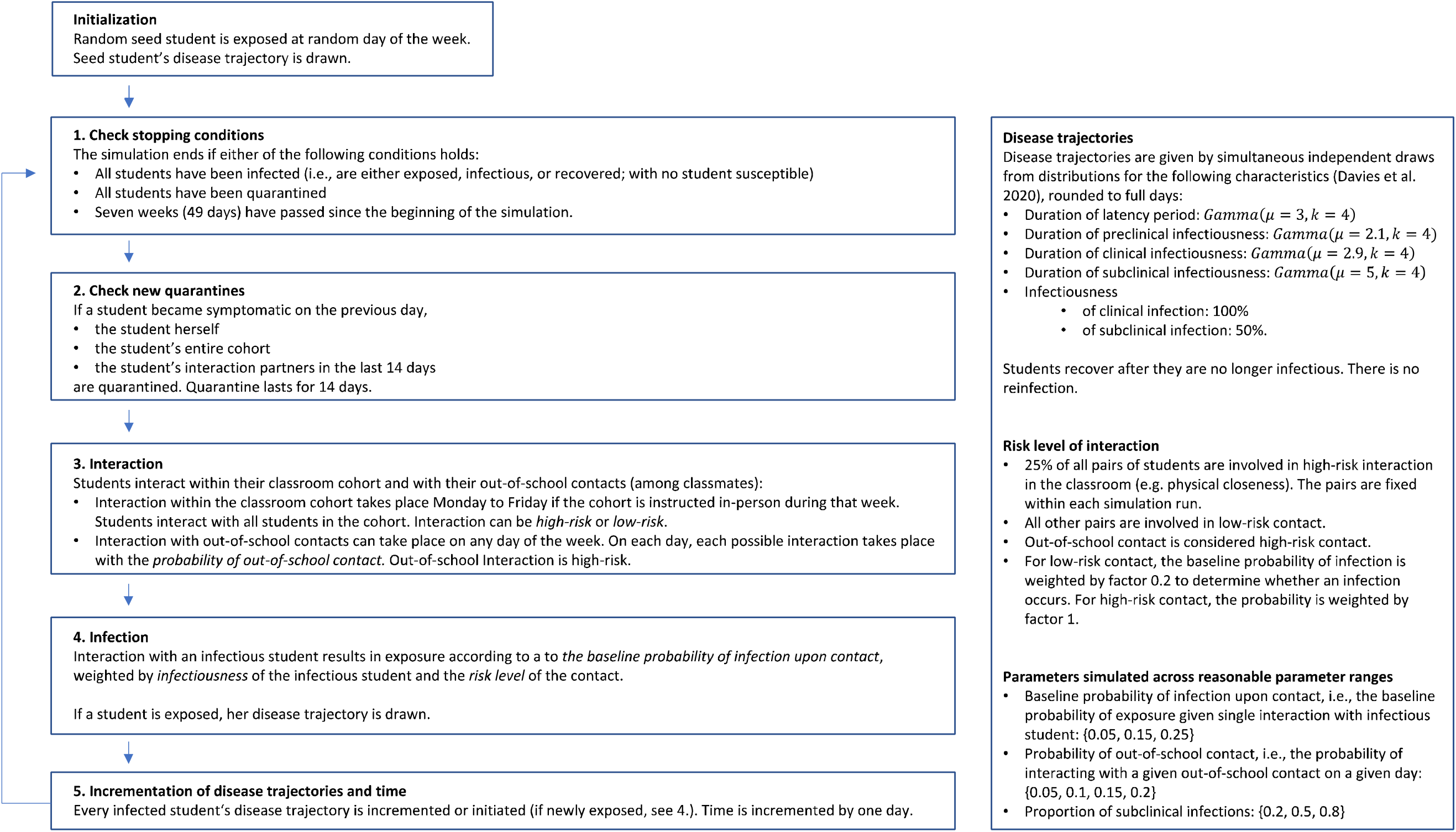
Model for transmission of SARS-CoV-2 within classroom

Each simulation starts with one randomly infected seed-node student who, once infectious, can infect her cohort members in school and her out-of-school contacts through out-of-school interaction. In-school interaction occurs only on school days (Monday to Friday) with in-person instruction, while out-of-school contact can occur every day of the week with a given probability. Similarly, contact with an infectious student results in infection with a given probability, so that infections are not deterministic either. When the seed node has infected additional students, they can in turn infect their cohort members and out-of-school contacts once they have become infectious.

Not all but some students have clinical infections. Once a student becomes symptomatic, we assume that all members of her cohort and all students involved in her out-of-school interactions in the last 14 days are quarantined on the next day according to a strict test-and-trace protocol. Quarantine lasts for 14 days.

Our agent-based model depends on a number of parameters. For students’ individual trajectories of Covid-19, we consider whether the infection is subclinical or clinical, the infectiousness of subclinical relative to clinical infection, the length of the latency period, the length of the infectious period, and the time until symptom onset given a clinical infection. We use the parameters from Davies et al.^9^ on these characteristics to stochastically model disease trajectories. These assumptions are themselves largely based on an aggregation of previous studies on SARS-CoV-2 and also summarized in Figure 2. For the infectiousness of subclinical relative to clinical infections, we consider 50% as a baseline, following Davies et al. In the Supplementary Material, we show that the results are similar when considering a relative infectiousness of 30%, the lower bound suggested in the recent literature^8,9^.

We model variation in three additional parameters that so far are highly uncertain: The probability of infection upon contact with an infectious student, the proportion of subclinical infections, and the probability of out-of-school contact. We bound the probability of infection upon contact to 5%-25%, which corresponds to average in-class secondary attack rates of 3%-19%, thus capturing most of the variation reported in the literature^29,32,54–57^. Similarly, we consider a proportion of subclinical student cases between 20% and 80% to depict the wide range of estimates on clinical cases among adolescents from recent research^8,9,58–61^. Finally, we consider daily probabilities of 5% to 20% for each out-of-school contact. This corresponds to averages of 1.05-4.20 out-of-school interactions per week for the median student, who has three out-of-school contacts. We use this range to capture the fact that contact behavior is likely to vary with the prevailing incidence of SARS-CoV-2 and with different legal regulations at different times and in different countries.

Results on the general effectiveness of cohorting are likely to depend on the dynamics of SARS-CoV-2 transmission in classrooms, which in turn depend on the parameters discussed above. Transmission dynamics are high when the probability of infection is high and the proportion of subclinical infections is high, preventing early quarantine. Under these conditions, SARS-CoV-2 spreads more easily and cohorting thus is likely to become more important. We therefore show all of our results for three scenarios: One scenario with low transmission dynamics, characterized by a low probability for infection (5%) and a low proportion of subclinical cases (20%). We contrast this with a scenario with medium transmission dynamics (probability for infection = 15%, proportion of subclinical cases = 50%) and a scenario with high transmission dynamics (probability for infection = 25%, proportion of subclinical cases = 80%). In Extended Data Figures 1 and 2 in the Supplementary Material, we show results across all combinations of parameter values. Throughout the analysis, we show results for different probabilities of out-of-school contacts. While the frequency of out-of-school contacts also affects transmission dynamics more generally, it is particularly likely to produce differences *between* cohorting strategies because it shapes the prevalence of cross-cohort transmission channels.

## RESULTS

We first compare random to no cohorting and find that *random cohorting* substantially reduces the risk of SARS-CoV-2 transmission in classrooms, even though it ignores out-of-school interaction. Figure 3 illustrates the average proportion of infected students across all classrooms, comparing no cohorting to random cohorting for both same-day instruction (left panel of Figure 3) and in a weekly rota-system (right panel). Infections strongly depend on transmission dynamics, with a much larger proportion of students infected when transmission dynamics are high and very few infections when dynamics are low. Independent of whether transmission dynamics are low or high, random cohorting reduces infections by about 50% compared to no cohorting. As a comparison of the left and the right panel shows, instruction in a weekly rota-system further reduces infections by about 50% relative to same-day instruction because it ensures that transmission within cohorts can only take place every other week. This “natural quarantine” frequently prevents larger outbreaks early on.

**Figure 3:**
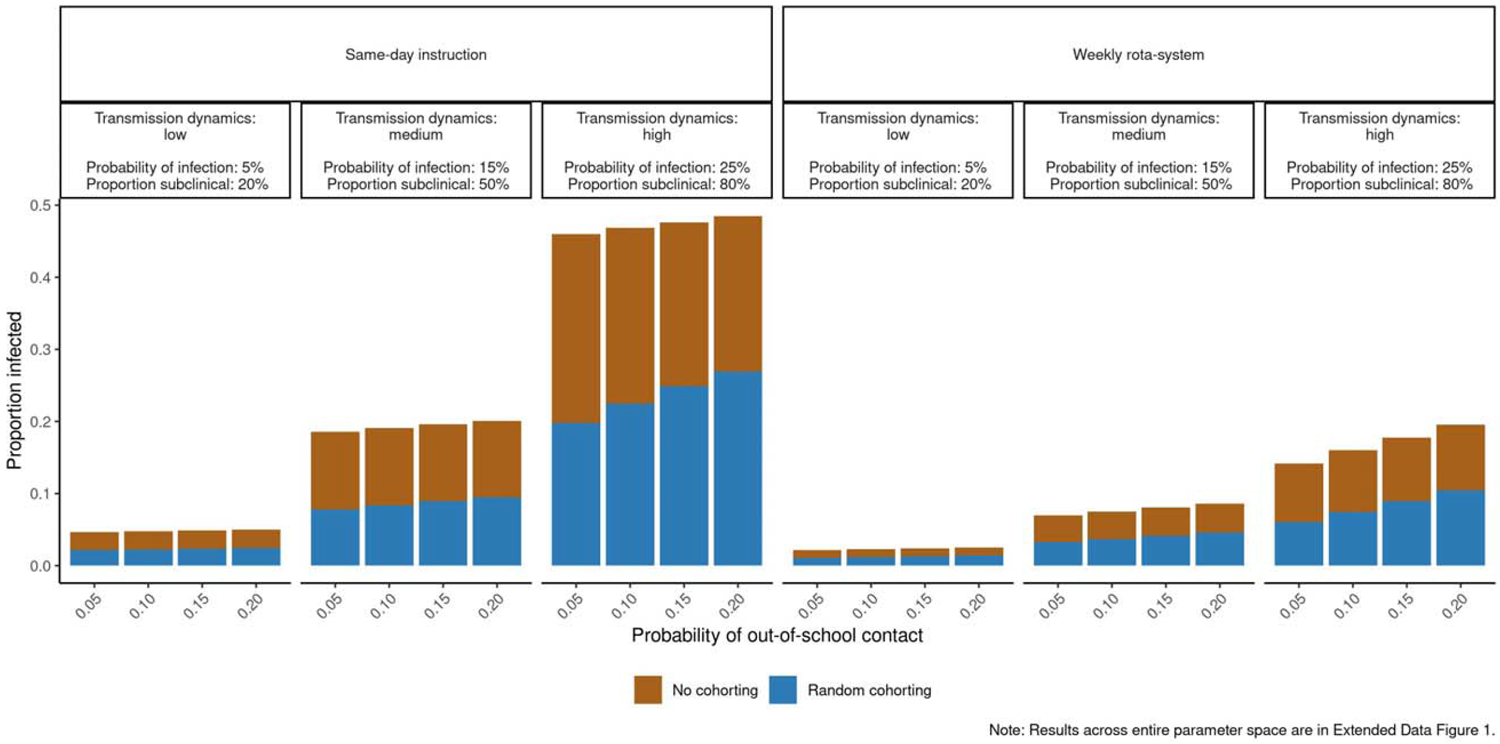
Average cumulative proportion of infected classroom members in case of random cohorting (blue) and no cohorting (brown + blue).

Figure 3 also shows that more frequent out-of-school contact increases infections, and this increase is similar in the non-cohorted classrooms and under random classroom cohorting. In relative terms, random cohorting therefore is particularly effective when out-of-school contacts are *infrequent*. When out-of-school contacts are more frequent, random cohorting becomes less effective because increasing cross-cohort infections cause additional outbreaks in the second cohort. This provides first indirect evidence that reducing the number of cross-cohort out-of-school contacts may help contain larger outbreaks.

Before turning to the simulation models for the social network-based cohorting strategies, Figure 4 shows the distribution of the average number of cross-cohort contacts across classrooms in all countries for the four different cohorting strategies described in Table 1. Across all countries, all cohorting strategies yield a number of cross-cohort ties that is substantively smaller than the total number of ties in the out-of-school network. As expected, the optimization strategy results in the lowest number of cross-cohort ties, with an average of 3.5 cross-cohort ties per classroom. This represents only 17% of the average of 20 cross-cohort ties resulting under random cohorting. The gender-split strategy produces an average of 11.4 cross-cohort ties, 57% of the cross-cohort ties under random cohorting. The network chain strategy results in an average of 8.4 cross-cohort ties, thus outperforming the gender-split strategy, with 42% of the cross-cohort ties under random cohorting remaining. Therefore, compared to the optimization strategy, network chain and gender-split cohorting are likely to be less efficient but to also help containing outbreaks.

**Figure 4:**
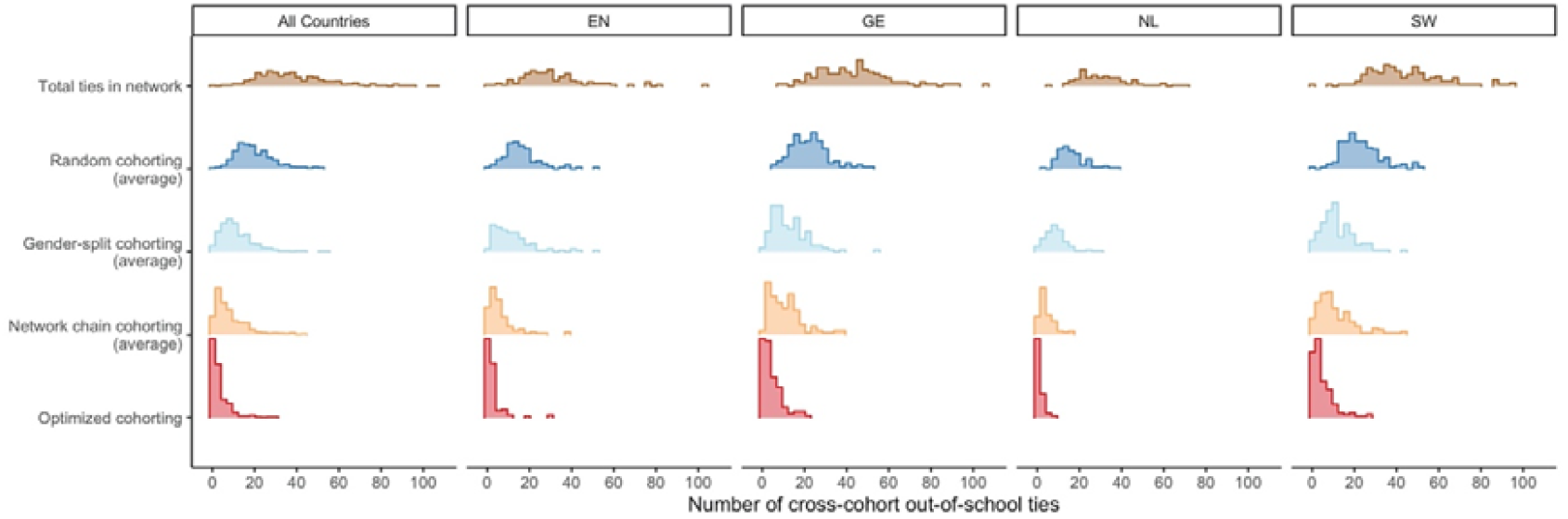
Total number of ties between classmates in the out-of-school contact network and (average) cross-cohort ties for different cohorting strategies across classrooms.

In our simulations, we show results for three indicators: the proportion of initial outbreaks that spread across cohorts, the proportion of infected students across the entire classroom, and the proportion of students quarantined (see Table 1). For quarantines, we only show the *excess proportion quarantined*: Any given proportion of clinical infections implies a specific minimum share of students who are quarantined independent of cohorting strategy. If, for example, 80% of all infections are clinical cases, 80% of seed nodes will eventually become symptomatic, triggering quarantine in their cohort and inducing a minimum of 40% of quarantined students on average. Therefore, we only show the excess proportion quarantined up and above this implied minimum share. Figure 5 shows results aggregated across the entire sample; classroom-level results by country are comparable and presented in the Supplementary Material.

**Figure 5:**
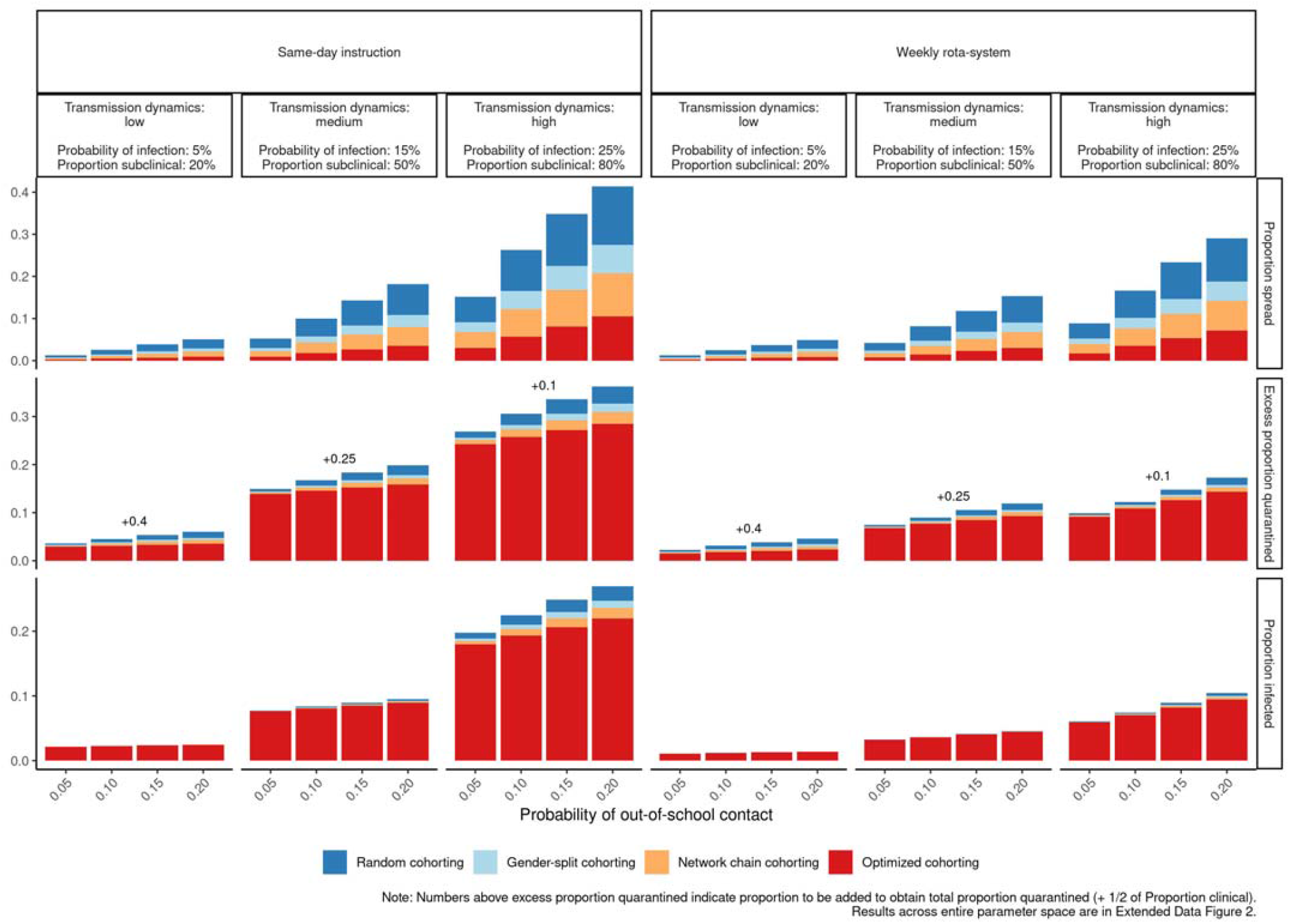
Epidemiological outcomes of different cohorting strategies: Proportion of instances of infection spreading to the second cohort, excess proportion of students quarantined, and proportion of students infected. Cumulative proportions.

The top row of Figure 5 shows that the frequency of SARS-CoV-2 spreading to the second cohort differs between cohorting strategies. Across all scenarios, gender-split, network chain and optimized cohorting all outperform random cohorting. Gender-split cohorting falls about halfway in between random and optimized cohorting. Network chain cohorting is more effective than gender-split cohorting but less effective than optimized cohorting. When transmission dynamics are stronger, infections of the second cohort are more frequent for all cohorting strategies. In addition, higher transmission dynamics exacerbate the differences between cohorting strategies, indicating that effective cohorting is particularly important when transmission dynamics are high. Throughout, weekly rota-systems buffer transmission to the second cohort relative to same-day instruction of both cohorts. More frequent out-of-school interaction increases the spread of SARS-CoV-2 to the second cohort and differences between the cohorting strategies grow further with rising probability of out-of-school contact. Effectively separating cohorts by avoiding cross-cohort out-of-school contact thus is particularly important when out-of-school contacts are frequent.

The general pattern of differences across cohorting strategies is similar for the excess proportion of students quarantined and the proportion of students infected across the entire classroom. In particular, optimized cohorting always performs best, followed by network chain cohorting, gender-split cohorting, and random cohorting. For two reasons, differences between the cohorting strategies are generally smaller for the proportion quarantined and infected than for the spread of SARS-CoV-2 to the second cohort. First, not every transmission of SARS-CoV-2 to the second cohort results in additional infections or quarantines. Second, most interactions (and transmissions) in the model occur within school rather than in less frequent out-of-school contact—which is the rationale behind cohorting in the first place. Therefore, a substantial baseline proportion of quarantines and infections are determined by within-cohort transmission dynamics.

Figure 5 shows that differences between cohorting strategies are small for these indicators when dynamics of transmissions are low. In particular, the total share of infections is almost independent of the cohorting strategy under low transmission because, under these conditions, outbreaks will die down quickly irrespective of the cohorting strategy. Differences between cohorting strategies also are small when a weekly rota-system is introduced. When cross-cohort transmission takes place, the rota-system frequently prevents onward transmission in the second cohort simply because the cohort only meets for in-person instruction every other week. However, when transmission dynamics are high, cohorting strategies that effectively prevent cross-cohort out-of-school contact also have substantive consequences for the overall proportion of infected students. When SARS-CoV-2 spreads to the second cohort under these conditions, onward transmission is likely, inducing a second outbreak that results in frequent infections and quarantines. Such additional outbreaks are less likely when cohorting strategies effectively prevent cross-cohort out-of-school contact.

Concerning epidemiological outcomes, an effective cohorting strategy thus is most important when transmission dynamics are relatively high. For example, at same-day instruction, a probability of out-of-school interaction of 20%, a baseline probability of infection upon contact of 15% and 80% subclinical infections, random cohorting on average results in 24% of outbreaks spreading to the second cohort, gender-split cohorting results in 15%, network chain cohorting in 11%, and optimized cohorting in 5%. Depending on its specific implementation, cohorting that considers out-of-school contact between classmates thus can lower the frequency of spread by 38%-78% relative to random cohorting. The excess proportion of quarantined students can be reduced from 22% (random cohorting) to 20% (gender-split), 19% (network chain cohorting) and 18% (optimized cohorting). Thus, excess quarantine can be reduced by 8-16% relative to random cohorting. The overall proportion quarantined is 10% higher because 20% of clinical infections imply a baseline proportion of quarantined students of 10%, but this baseline proportion cannot be affected by any specific cohorting strategy. The average proportion of infections at the same time falls from 13.7% (random cohorting) to about 12.9% in gender-split, 12.6% network chain and 12.1% in optimized cohorting. Relative to random cohorting, infections thus are reduced by 5.9% (gender-split strategy), 8.0% (network chain strategy), and 11.6% (optimized strategy), respectively. While these reductions appear modest in size, it is important to bear in mind that, especially in a situation with high incidence of SARS-CoV-2, they are likely to apply to a large number of classrooms and can thus prevent a large aggregate number of infections and quarantines. Furthermore, other than cohorting *itself*, applying a specific cohorting *strategy* does not come with large organizational costs or reduced in-person instruction but only requires allocating cohorts in a specific way.

Substantively, the usefulness of cohorting strategies that prevent cross-cohort out-of-school interactions thus depends both on how easily SARS-CoV-2 is transmitted among students and the underlying goals of cohorting. If transmission is low, the main advantage of effective cohorting is to reduce the frequency of quarantines, thus keeping students in school more. When transmission is more dynamic—at higher probabilities for infection or a higher share of subclinical cases—effective cohorting also reduces the total burden of infections notably by frequently containing larger outbreaks to a single cohort rather than allowing them to spread to the second cohort. This also holds true when the probability of out-of-school interaction is high, which more frequently induces a second outbreak after containment has failed in one cohort.

## DISCUSSION

At the turn of the year 2020/2021, the SARS-CoV-2 pandemic still is a disruptive force in many areas of society. Social distancing measures, unprecedented in scope, have helped mitigate the first wave of infections, though at high costs. Among these high-cost measures were school closures, which resulted both in missed learning opportunities for children and in considerable strain on their families. School closures therefore are seen as a last resort when other measures have failed or were insufficient to prevent the spread of SARS-CoV-2. With increasing incidence of SARS-CoV-2, however, infections in schools become more likely, requiring effective social distancing strategies to avoid transmission and larger outbreaks in schools.

One such strategy is cohorting, the decomposition of larger clusters of students into smaller isolated units. In the European and other contexts where students are taught in classrooms of 20 to 40 students, cohorting strategies are best applicable to splitting classrooms in half. Simulating the transmission of SARS-CoV-2 in classrooms and out-of-school contact networks of students in England, Germany, the Netherlands, and Sweden, we show that cohorting helps contain outbreaks, substantially reducing the number of infected students. It proves particularly effective when conducted in a rota-system, with each cohort receiving in-person and remote instruction in alternating weeks. Cohorting is successful because it facilitates social distancing and helps contain initial outbreaks in a single cluster. Combining cohorting with a rota-system is even more effective because infectious students cannot transmit the disease in the classroom in the remote learning weeks, halting outbreaks or preventing them in the first place.

However, the success of cohorting depends on whether cohorts can be isolated not only *within* the school context, but also in terms of *out-of-school* interaction. Therefore, we compare random cohorting, which does not consider out-of-school contact, with three network-based cohorting strategies: gender split cohorting, optimized cohorting, and network chain cohorting (see Table 1). These network-based strategies account for students’ out-of-school contact with classmates in allocating them to cohorts, trying to reduce cross-cohort out-of-school contacts. All of them outperform random cohorting by more frequently containing outbreaks to a single cohort. They also reduce the frequency of quarantines and the number of students infected, though the latter effects are weaker when transmission dynamics are limited—i.e., when instruction is organized in a rota-system, the risk of infection is low, the share of clinical cases is high (inducing early quarantine), and/or out-of-school contact is rare. In this case, their main benefit is to produce fewer quarantines, thus keeping students in school more.

The fact that splitting cohorts by gender reduces transmission dynamics reflects that adolescents’ out-of-school contacts are mostly among students of the same gender. Optimized cohorting proves even more effective, because it explicitly minimizes the number of cross-cohort out-of-school contacts. However, since this strategy requires full knowledge of students’ out-of-school contact with classmates, it might be difficult to implement in practice. Fortunately, network chain cohorting offers a simple approximation which also performs better than random allocation and gender-split cohorting. In this strategy, an initial student names all classmates she meets outside of school. The nominated students in turn indicate their within-class out-of-school contacts until the resulting nominations comprise half of the class. The resulting set of students then constitutes one cohort, the remainder of students the other. While less effective than explicit optimization, this strategy is easier to implement in practice, as schools do not need full information on contact networks but can allocate students through a simple method. Network chain cohorting thus offers a good compromise between effectiveness and practicability.

From a pedagogical viewpoint, some of our cohorting strategies may have drawbacks. For example, network chain cohorting may cause socially awkward situations because only some students are asked to name their out-of-school contacts, because some students may be disappointed when not nominated, or because students may refuse to cooperate. By design, however, this strategy protects isolated students from being publicly exposed in the classroom as an entire half of the classroom, rather than only isolated students, is not nominated. Splitting classrooms by gender may also be undesirable, especially in cases where some students have to be allocated to the other-gender cohort because of gender imbalance in the classroom. Teachers, school administrators, and policy makers need to weigh these potential pedagogical drawbacks against the benefits of each strategy and make decisions accordingly.

Irrespective of pedagogical concerns, gender-split cohorting also has an epidemiological downside that our model does not address. If students maintain heterosexual romantic relationships, they carry an elevated transmission risk and may thus serve as infection channels between gender-specific cohorts. Separating cohorts by gender therefore may backfire by opening some particularly transmission-prone cross-cohort channels.

Our model might furthermore miss epidemiologically relevant processes because we do not model the full spectrum of students’ interaction and transmission channels. For instance, we do not consider teachers, who may carry the disease from cohort to cohort. However, because teachers’ impact on transmission between cohorts is constant across our strategies, the relative evaluation of the cohorting strategies should remain unaffected. Similarly, the out-of-school contacts we consider are limited to classmates and do not extend to parents or siblings. In reality, if an infected student induces a symptomatic infection in such an interaction partner, this may trigger a (delayed) quarantine in the classroom when the student is tested belatedly. However, this is also unlikely to change conclusions about the relative effectiveness of the different cohorting strategies.

As all agent-based models, our model rests on core assumptions, and changes in these assumptions could change the outcomes we observe. First, all strategies we discussed assume that some classmates meet outside of school. While our model accounts for considerable reductions in contacts during the pandemic, we do not consider a complete halt of out-of-school contacts. In a very strict lockdown scenario without any out-of-school contact, allocation to cohorts according to contact networks becomes irrelevant or could even be harmful under certain conditions. If there is no transmission between cohorts because students cease to meet after school, the focus will shift to infection probabilities within classrooms. If the risk of infection in the classroom is elevated between students who meet after school, it may be beneficial to allocate close contacts to different rather than identical cohorts. Second, in our model, symptomatic students are quickly tested and quarantined. If high local incidence of SARS-CoV-2 leads to delays in testing or quarantines, effective cohorting becomes more important. Third, our model is sensitive to changes in parameters that rely on current imperfect knowledge about SARS-CoV-2. New data on the role of adolescents in the transmission of SARS-CoV-2 may change modeling assumptions. We investigate plausible ranges for all relevant parameters, and model results vary across the parameter space, with weaker effects of cohorting strategies when transmission dynamics are low.

A strength of our study is the use of complete empirically observed classroom network data instead of ego-centered or synthetic networks. As a consequence, we are able to capture the clustering patterns of real-world networks and their implications for social distancing strategies. As a downside, school-based network survey data are often incomplete, as students might forget to nominate classmates or restrict their nominations because they want to complete the survey more quickly. It is also important to note that our data is from 2010-2011, and interaction patterns among students may have changed. While there is no immediate reason to expect these limitations affect our qualitative conclusions, they indicate that future research is needed both on the physical propensities of adolescents to transmit SARS-CoV-2 and the behavioral patterns of this age group under pandemic conditions.

In sum, our study shows that cohorting can decrease the transmission of SARS-CoV-2 in classrooms. The way in which classrooms are divided matters. We have demonstrated that simple and easily implementable network-based strategies can improve the effectiveness of cohorting by reducing cross-cohort out-of-school interaction with classmates. The ensuing separation between cohorts limits the spread of SARS-CoV-2 across cohorts and can further reduce quarantines and infections, especially in situations with strong transmission dynamics.

## METHODS

### The CILS4EU data

Our simulations use data from the first wave of the Children of Immigrants Longitudinal Study in Four European Countries (CILS4EU) project^51,52^, which provides information on 14-15-year-old students from England, Germany, the Netherlands, and Sweden. Within each country, data was collected in 2010-11 in randomly selected schools, oversampling schools with a high share of immigrant students. In most schools, two ninth-grade classrooms were surveyed in full, providing individual student information as well as data on social relations between surveyed students within a classroom. The response rate at the student level was 81% in England and Germany, 91% in the Netherlands, and 86% in Sweden^62^. Our analysis considers all classrooms with information on 20 or more students because cohorting is likely to be less of an issue in small classes. To compare the gender-split strategy with other strategies, we only consider classrooms with full information on students’ gender. The resulting sample consists of 507 classrooms populated by 12,291 students.

Our indicator of out-of-school contacts considers all classmates that a focal student indicated to “often spend[s] time with outside school”. Students could nominate as many of their classmates as they wanted. In many classrooms, however, students were only allowed to nominate students who also participated in the survey. Therefore, we limit all our networks to students who participated in the student and/or network questionnaire. Whenever one student named another, we code an out-of-school contact between this pair of students, independent of whether the second student confirmed the nomination, because contact necessarily goes both ways.

### Implementation of cohorting strategies

In our analyses, we consider four cohorting strategies (see Table 1 for a summary) and here, we provide technical details on the implementation of these strategies. For *random cohorting*, each classroom is randomly split into two equally-sized cohorts. If the number of students is odd, one cohort exceeds the other in size by one student.

In the *gender-split* cohorting strategy, cohorts are separated by gender. If a strict separation of boys from girls leads to unequal cohort sizes because of an uneven gender composition, members of the larger cohort (the overrepresented gender) are reallocated to the smaller cohort until cohort sizes equalize. There are also other approaches to forming cohorts by gender if gender representation is unequal, two of which are explored in the Supplementary Material. However, as explained in the Supplementary Material, ensuring equal group sizes is preferable from the perspective of preventing infections and quarantine.

In the *optimized* cohorting strategy, students are allocated to cohorts in order to minimize the number of cross-cohort out-of-school contacts with classmates. We use brute-force optimization, considering all possible allocations to equally-sized cohorts to find the minimum number of cross-cohort out-of-school contacts. In eight classrooms with more than 32 students, optimization fails due to computational constraints. For these classrooms, we randomly sample 1,000,000 allocations and report results for the allocation that minimizes the number of cross-cohort out-of-school contacts.

*Network chain* cohorting uses chains of out-of-school contact nominations to allocate students to cohorts. In this strategy, a random well-connected student names all of her out-of-school contacts and this set of students forms the core of the first cohort. In the simulations, we draw the initial student from the observed out-of-school relations, with probability proportional to the number of out-of-school contacts. Therefore, better-connected students are more likely to be selected as initial students. This simplifies allocation because the algorithm is less likely to break down. The nominated out-of-school contacts themselves subsequently nominate their (not yet nominated) out-of-school contacts, continuing this process until the set of nominated students comprises half of the classroom. This set forms one cohort and the remaining students are pooled in the second cohort. If there are no additional nominations at a certain nomination step and the set does not yet comprise half of the students, a random student is added to the set and can subsequently nominate her contacts. If, during the nomination process, the number of students in the group would exceed half of the class size due to new nominations, a random subset of the newly-nominated students is added to the set. These rules ensure that the algorithm always ends up with a definitive allocation.

### Setup of agent-based simulation models of transmission dynamics in classrooms

To simulate SARS-CoV-2 outbreaks, we run agent-based models for each of the 507 classrooms and for each cohorting strategy. Given that simulation outcomes are stochastic, we run 2000 simulations for each cohorting strategy in each classroom. Each simulation run starts with one random infected student. The simulation then assesses how SARS-CoV-2 spreads within the classroom from this seed node. The simulation ends when all students have been infected or quarantined, or when seven weeks have passed, capturing the effect of school holidays. Figure 2 gives an overview of the simulation model.

In the simulation model, infections stem from *interaction* with an infectious classmate. Interaction is modelled on a daily basis and can occur both within and outside of school. In-school interaction occurs with all cohort members on school days (i.e., Monday to Friday) and only when the cohort is instructed in-person (i.e., every other week when cohorts are instructed in a rota-system). We model differential risks of infection in the classroom context to capture the effects of physical proximity, assuming a high risk of infection for a randomly chosen 25% of the within-cohort pairs of students. As a cohort usually consists of 10-16 students, this results in an average of 2-4 high-risk interactions, which arguably resembles the number of students who will be in close physical proximity within a cohort. For the remaining students, we assume a lower risk (20% of the high-risk contact), capturing the effect of aerosol diffusion and other possible transmission routes such as movement in breaks and possibly fomite transmission. We also assessed 12.5% and 50% high-risk contacts, with similar qualitative results but different baseline transmission of SARS-CoV-2 in the classrooms, as documented in the Supplementary Material.

Out-of-school interaction can occur on any day of the week. Because out-of-school contact is likely to entail intense and enduring interaction, we assume out-of-school contact to have the same risk of infection as high-risk contact within the classroom. We model out-of-school contact probabilistically, with a given and independent probability of any out-of-school contact taking place on any day.

Irrespective of whether interaction with an infectious student occurs in school or outside of school and whether it is a high-risk contact or not, it does not lead to infection in all cases. We model the likelihood of infection with a baseline probability of infection upon contact, as discussed below. This baseline probability is further modified by the differential infection risk in the classroom and the infectious student’s infectiousness. Once a student (either through the seed node or through longer chains of infection) becomes infectious herself, she can infect other students.

Some, but not all, students have clinical cases. Once a student becomes symptomatic, all members of her cohort and all students involved in her out-of-school interactions in the last 14 days are quarantined one day later. Quarantine lasts for 14 days in which none of the quarantined students has contact with any classmate.

To model infections among students, we simulate their Covid-19 disease trajectories. We characterize whether an infection is subclinical or clinical, the infectiousness of subclinical and clinical infection, the latency period, the length of the infectious period, and the time until symptom onset given a clinical infection. We use the same parameters as Davies et al.^9^ on these characteristics of disease trajectories (summarized in Figure 2), which are themselves largely based on an aggregation of previous studies on Covid-19. Disease trajectories are stochastic, with the length of the latency period (mean: 3 days), the length of the infectious period (mean: 5 days), and the time until symptom onset after the infectious period has started (mean: 2.1 days) drawn from Gamma distributions (see Figure 2) and rounded to full days each. Along the lines of Davies et al., we assume that the infectiousness of subclinical cases is 50% of the infectiousness of clinical cases. As documented in the Supplementary Material, our findings are similar when we consider a relative infectiousness of 30%, which are the lowest estimates in the literature^8^.

### Model parameters

While good estimates for several characteristics of Covid-19 trajectories are available (see preceding paragraph), other key model parameters are so far unknown, highly uncertain, or context-specific. This especially holds true for the probability of infection conditional on contact with an infectious student, the probability of out-of-school interaction, and the proportion of clinical infections with SARS-CoV-2.

Naturally, model outcomes are sensitive to the specification of these parameters. At a higher likelihood of infection, larger outbreaks are more likely; at higher probabilities of out-of-school interaction, out-of-school contacts become more relevant; a higher share of clinical infections triggers quarantines more quickly, but symptomatic cases may also be more infectious relative to asymptomatic infections. Lacking precise estimates for these parameters, we model results for a range of plausible values, as discussed subsequently.

### Baseline probability of infection upon contact

The probability of infection upon contact is contingent both on adolescents’ general susceptibility to SARS-CoV-2, which is still unclear^29,32,63^, and on how conducive the school context is for transmission. To some degree, classrooms provide ideal conditions for transmission as they are small, confined indoor spaces that people share for multiple hours. However, during the SARS-CoV-2 pandemic, many schools have adapted to pandemic conditions by enforcing frequent ventilation, the usage of masks, and other precautions.

We simulate model results across a range of infection probabilities. We model a *baseline probability of infection upon contact*, which is the probability of becoming infected conditional on exposure to an infectious student on any given day. Same-day interaction within school and outside of school are considered to be separate encounters that pose distinct infection risks. It is a *baseline* probability because this probability is further reduced if it involves a low-risk contact and is modified by the infectiousness of the infectious student. We consider baseline probabilities of 5%, 15%, and 25%. From these probabilities, we can calculate corresponding in-classroom secondary attack rates for average seed nodes, considering the number of days they are infectious, how likely each day is to be a school day, and their classmates’ infection risks from an interaction. For the average seed node, a baseline probability of 5% corresponds to an in-classroom secondary attack rate of 3% for subclinical and 4% for clinical cases. A baseline probability of 25% represents a secondary attack rate of 16% for subclinical and 19% for clinical cases. This corresponds well with the range of estimates for the secondary attack rate among adolescents from previous studies ^29,32,55–57^. While Dattner et al. report a household secondary attack rate among 7 to 19-year-olds of 34%^in 32^, thus exceeding our upper bound, this is an outlier. We do not consider baseline probabilities of infection that exceed 25% because, in our model, these frequently result in very large classroom outbreaks, which we would expect to be able to observe in the real world and thus seem less plausible given the other model parameters.

### Probability of out-of-school interaction

For each out-of-school contact we model a fixed, independent, daily probability of interaction. Realistic values for this parameter will vary with conditions in society. With a high incidence of SARS-CoV-2 or a lockdown that prohibits certain contacts, probabilities for interaction will be lower than under normal circumstances. We consider probabilities of students to meet an out-of-school contact on a given day of 5%, 10%, 15%, and 20%. In our data, the median student nominates three (mean of 3.58) classmates who she has frequent out-of-school contact with. A probability of out-of-school contact of 5% thus means that the median student on average has out-of-school interactions in a week, with a probability of 86% to not have any out-of-school interaction on any given day and a probability of 34% to not have any interaction in an entire week. A probability of out-of-school contact of 20% means that the median student has on average 4.2 out-of-school interactions per week, with a probability of 51% of having no contact on any given day and a probability of just below 1% of having no contact in an entire week. This should capture a range of plausible contact frequencies under different conditions of the pandemic.

### Proportion of subclinical infections among adolescents

The proportion of subclinical SARS-CoV-2 infections among children and adolescents is highly debated. Even if the proportion of truly asymptomatic infections is low, many other infections may come with very weak symptoms and thus go undiagnosed, especially in high-incidence situations with limited access to testing and in seasons where cold- or flu-like symptoms are widespread. Previous studies suggest a wide range of estimates for the share of subclinical infections, ranging from 22%^59^ to 86.6%^8^ among children and adolescents with other estimates in between^9,58,60,61^. To capture the high uncertainty in this parameter, we model transmission processeh with assumed proportions of subclinical infections of 20%, 50%, and 80%.

## Data Availability

Data from: Kalter, Frank, Anthony F. Heath, Miles Hewstone, Jan O. Jonsson, Matthijs Kalmijn, Irena Kogan, and Frank van Tubergen. 2016. Children of Immigrants Longitudinal Survey in Four European Countries (CILS4EU)-Full Version. Data File for On-site Use. doi:10.4232/cils4eu.5353.1.2.0.

## EXTENDED DATA FIGURES

**Extended Data Figure 1:**
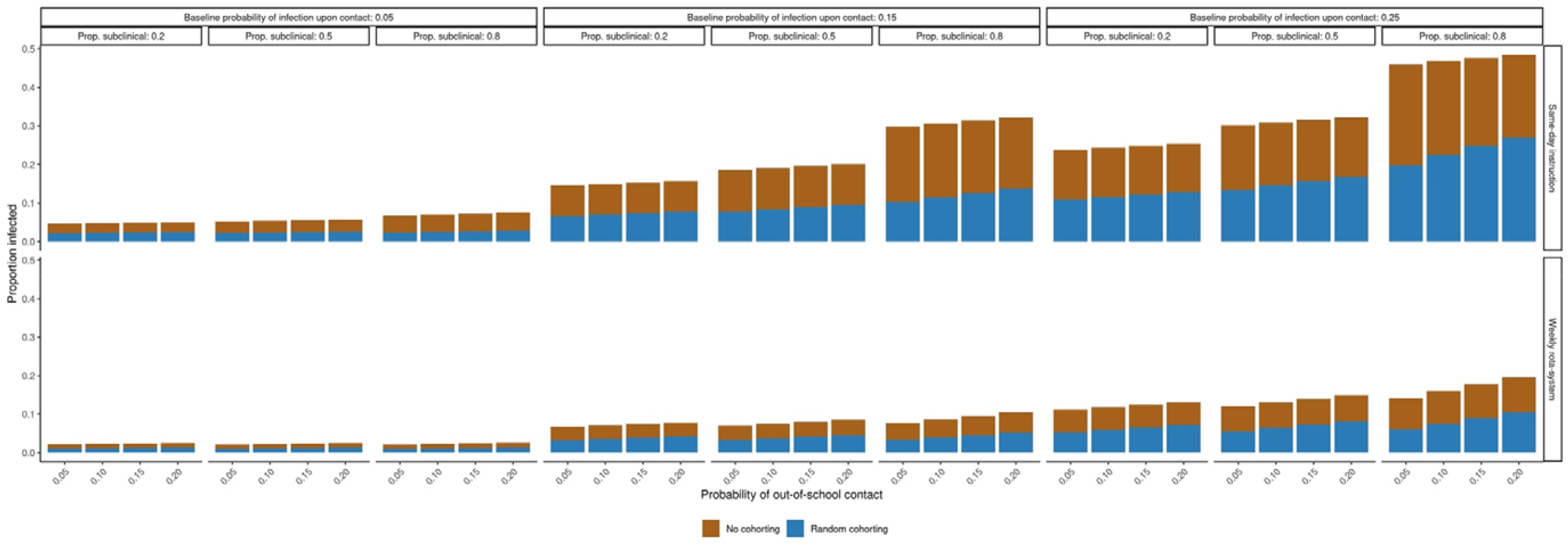
Results across the entire parameter space. Average cumulative proportion of infected classroom members in case of random cohorting (blue) and no cohorting (brown + blue)

**Extended Data Figure 2:**
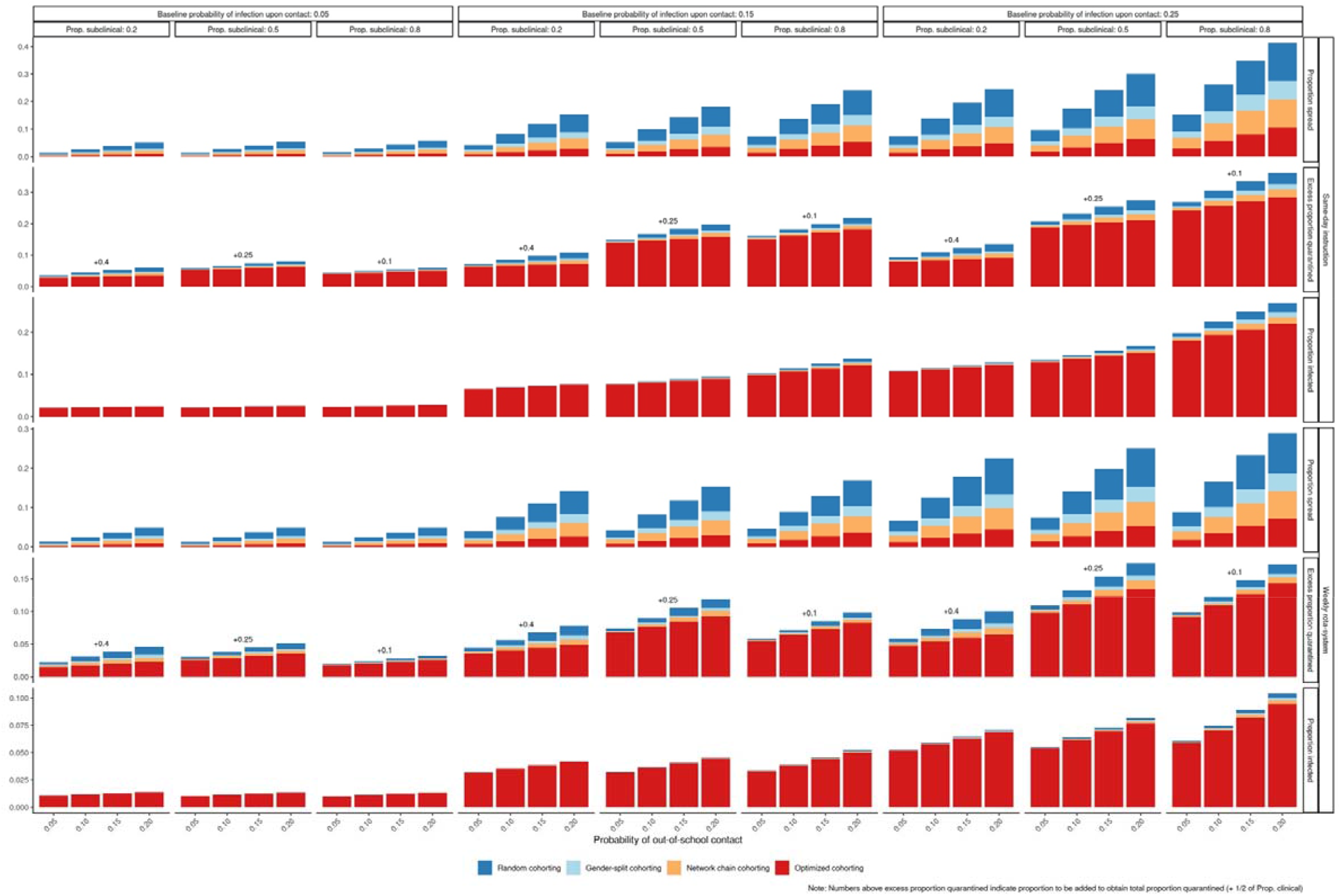
Results across the entire parameter space. Epidemiological outcomes of different cohorting strategies: Proportion of instances of infection spreading to the second cohort, proportion of students infected, and excess proportion of students quarantined. Cumulative proportions.

