## Supplemental Information for "Social network-based strategies for classroom size reduction can help limit outbreaks of SARS-CoV-2 in high schools. A simulation study in classrooms of four European countries"

**Supplementary Material**

Index:


A. Comparing variants of the gender-split strategy
B. Country- and classroom-specific results: Comparison of no cohorting to random cohorting and different cohorting strategies
C. Epidemiological outcomes for lower infectiousness of subclinical relative to clinical infections
D. Epidemiological outcomes for different proportions of in-classroom high-risk interactions

### A. Comparing variants of the gender-split strategy

In the main text, we focus on gender-split cohorting that enforces identical cohort size by reallocating students from the overrepresented gender group to the other cohort. There are other ways to deal with gender imbalances in classrooms. In this supplementary material, we compare the strict reallocation strategy from the main text both to a strategy without reallocations (i.e., accepting different cohort sizes) and to a strategy that only performs reallocations in the case of substantially different cohort sizes. In this latter strategy, we consider a difference in cohort size of five or more students as a cutoff. The rationale for not enforcing identical cohort size in these situations is that the reallocation of a student to the cross-gender cohort is likely to produce many cross-cohort out-of-school contacts because contacts are concentrated among students of the same gender. This can also be seen from Figure A1, which shows that the number of cross-cohort out-of-school contacts is highest in the gender-split strategy that enforces equal cohort size and lowest in the gender-split strategy that tolerates all cohort size differences. Therefore, the transmission of SARS-CoV-2 to the other cohort is likely to be more frequent when enforcing equal cohort size.

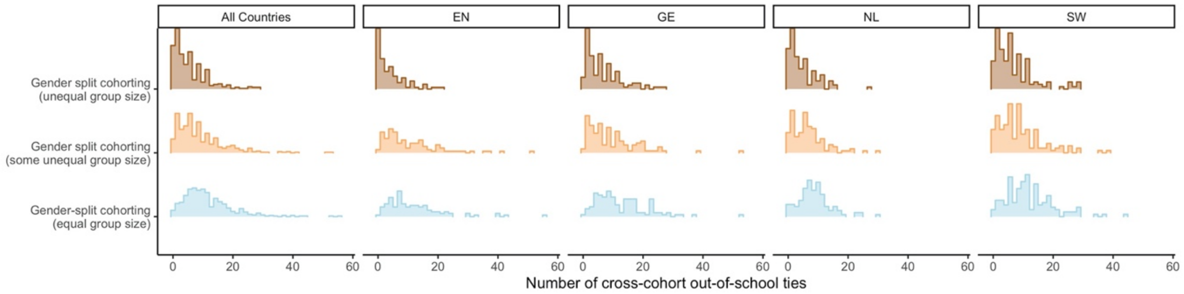


**Figure A1:** (Average) number of cross-cohort ties for different variants of gender-split cohorting across classrooms.

However, unequal cohort size can also have negative epidemiological consequences. Cohort size is likely to be exponentially rather than linearly related to transmission dynamics. When cohorts are unequal in size, the stronger transmission dynamics in the larger cohort are therefore likely to overcompensate the weaker transmission dynamics in the smaller cohort. In Figure A2, we show epidemiological outcomes from simulations for the different variants of the gender-split strategy to assess whether the effect of unequal cohort size is stronger than the effect of more frequent cross-cohort out-of-school contacts.

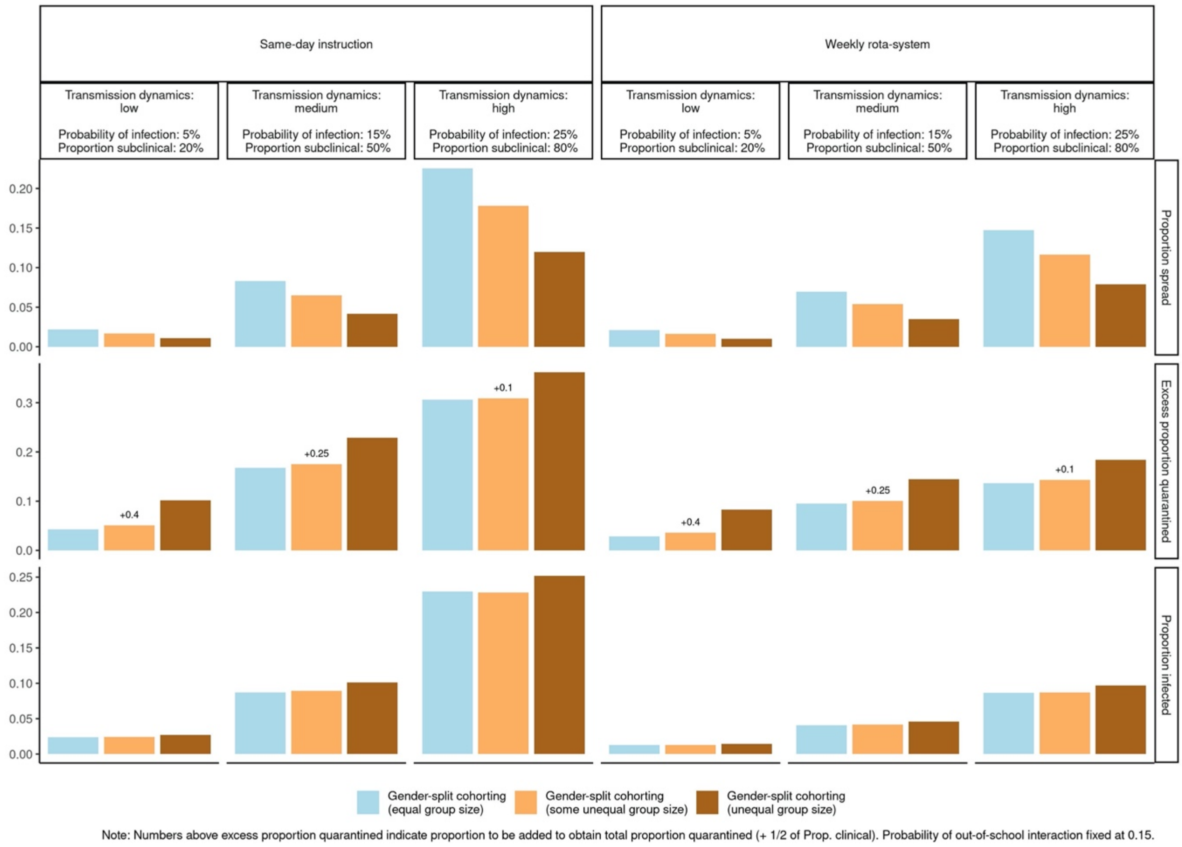


**Figure A2:** Epidemiological outcomes of different gender-split cohorting strategies: Proportion of instances of infection spreading to the second cohort, proportion of students infected, and excess proportion of students quarantined. Cumulative proportions.

As expected, Figure A2 shows fewer instances of transmission to the second cohort when different cohort sizes are allowed, and most strongly so when there are no limits to differences in cohort size. This pattern, however, is not mirrored in the total proportion of students infected or quarantined, which are both higher when unequal cohort sizes are allowed, in particular, when there are no restrictions on cohort size differences. This suggests that, at least for the parameter ranges we consider, the effect of unequal cohort size dominates the effect of reduced cross-cohort out-of-school contact. At least in terms of epidemiological outcomes, reallocating students to ensure equal cohort size thus seems preferable, which is why we focus on this variant of the gender-split strategy in the main text.

### B. Country- and classroom-specific results: Comparison of no cohorting to random cohorting (Fig. B1) and different cohorting strategies (Fig. B2-B4)

Each of the following figures shows epidemiological outcomes separately for each classroom, sorted by country. Classrooms are displayed on the x-axis. For each panel with a specific probability of infection and proportion of subclinical cases, classrooms are ordered by country, with country abbreviations printed inside the figures. Dotted vertical lines separate classrooms from different countries.

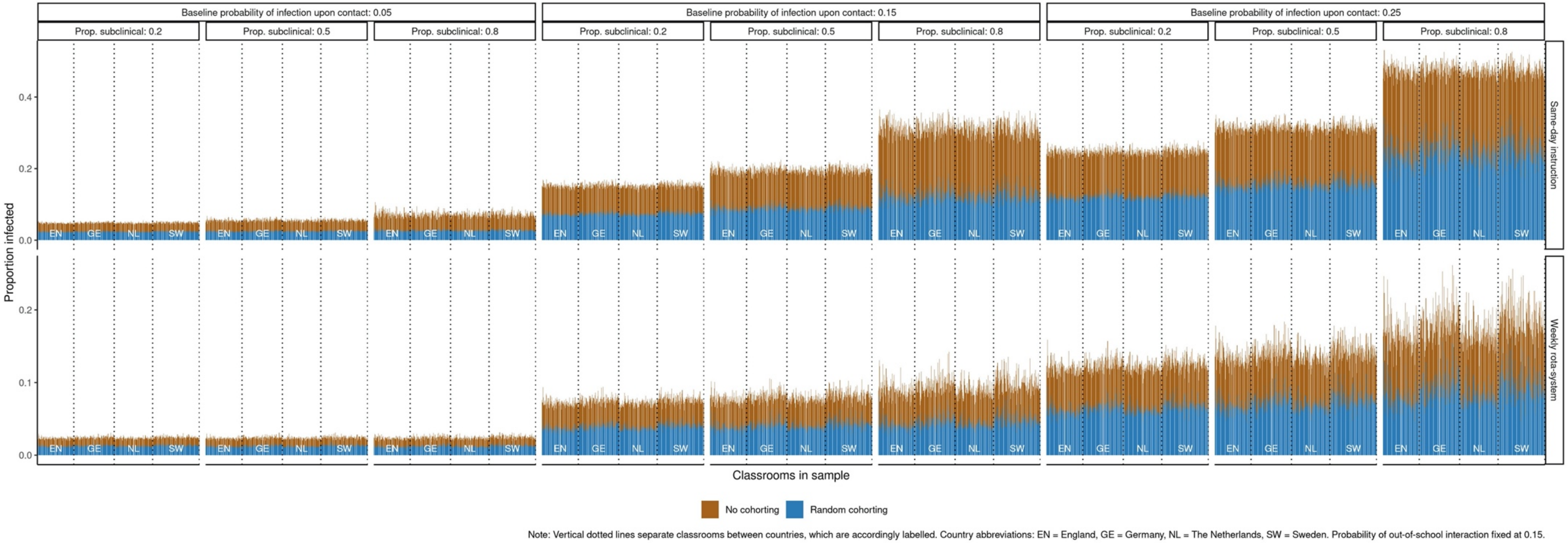


**Figure B1:** Average cumulative proportion of infected classroom members in case of random cohorting (blue) and no cohorting (brown + blue) for each classroom.

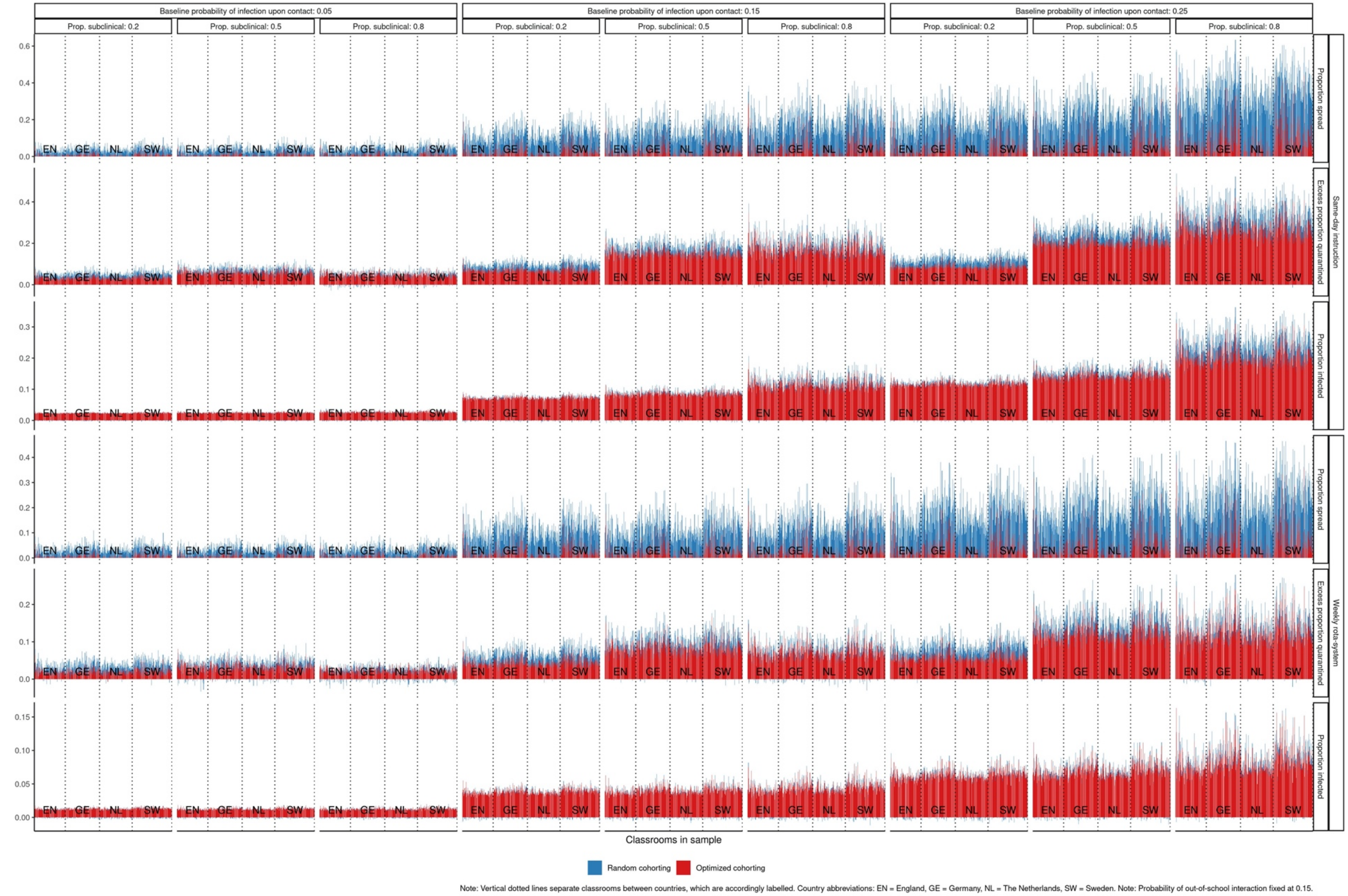


**Figure B2:** Random and optimized cohorting strategies. Epidemiological outcomes for each classroom: Proportion of instances of infection spreading to the second cohort, proportion of students infected, and excess proportion of students quarantined. Cumulative proportions.

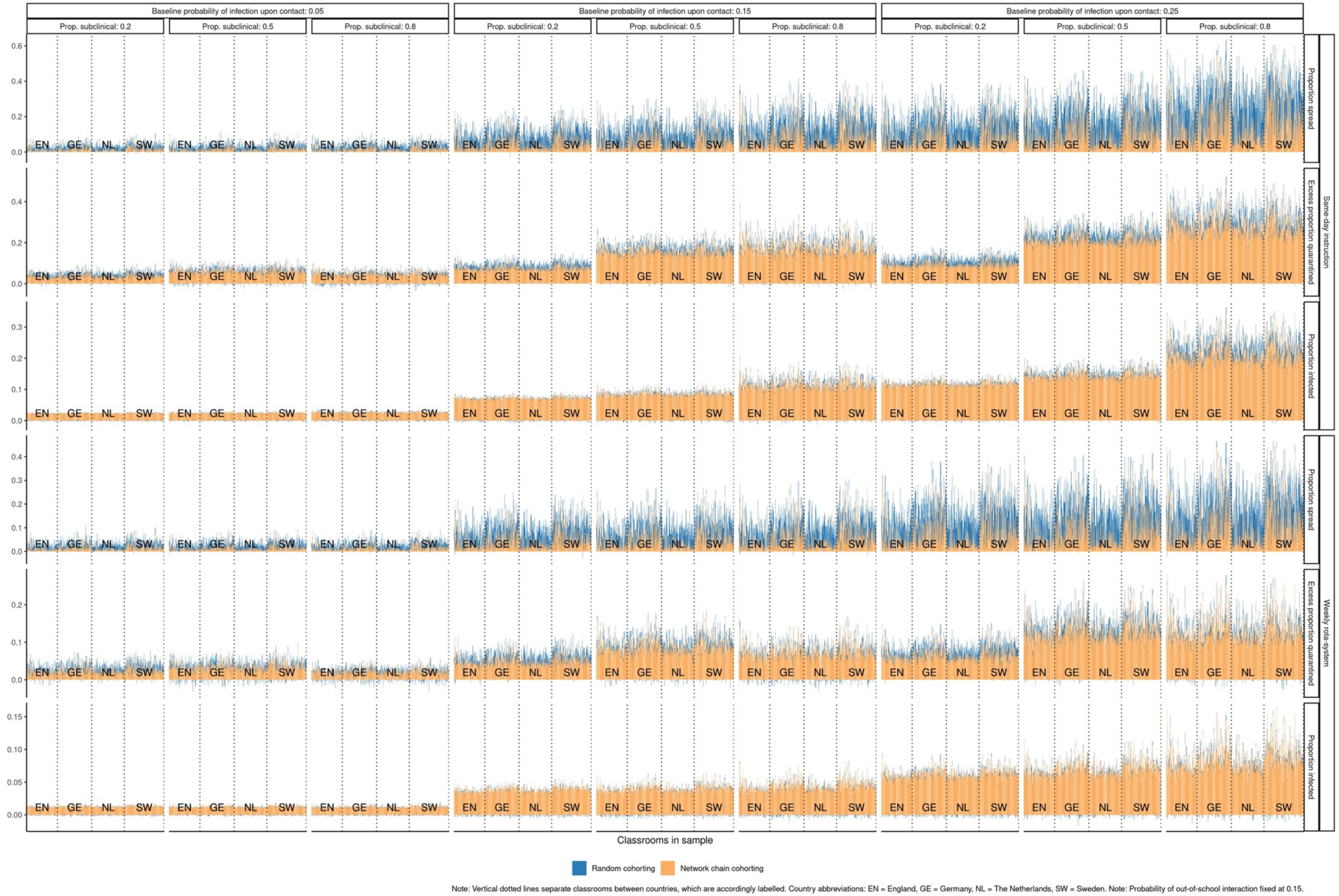


**Figure B3:** Random and network chain cohorting strategies. Epidemiological outcomes for each classroom: Proportion of instances of infection spreading to the second cohort, proportion of students infected, and excess proportion of students quarantined. Cumulative proportions.

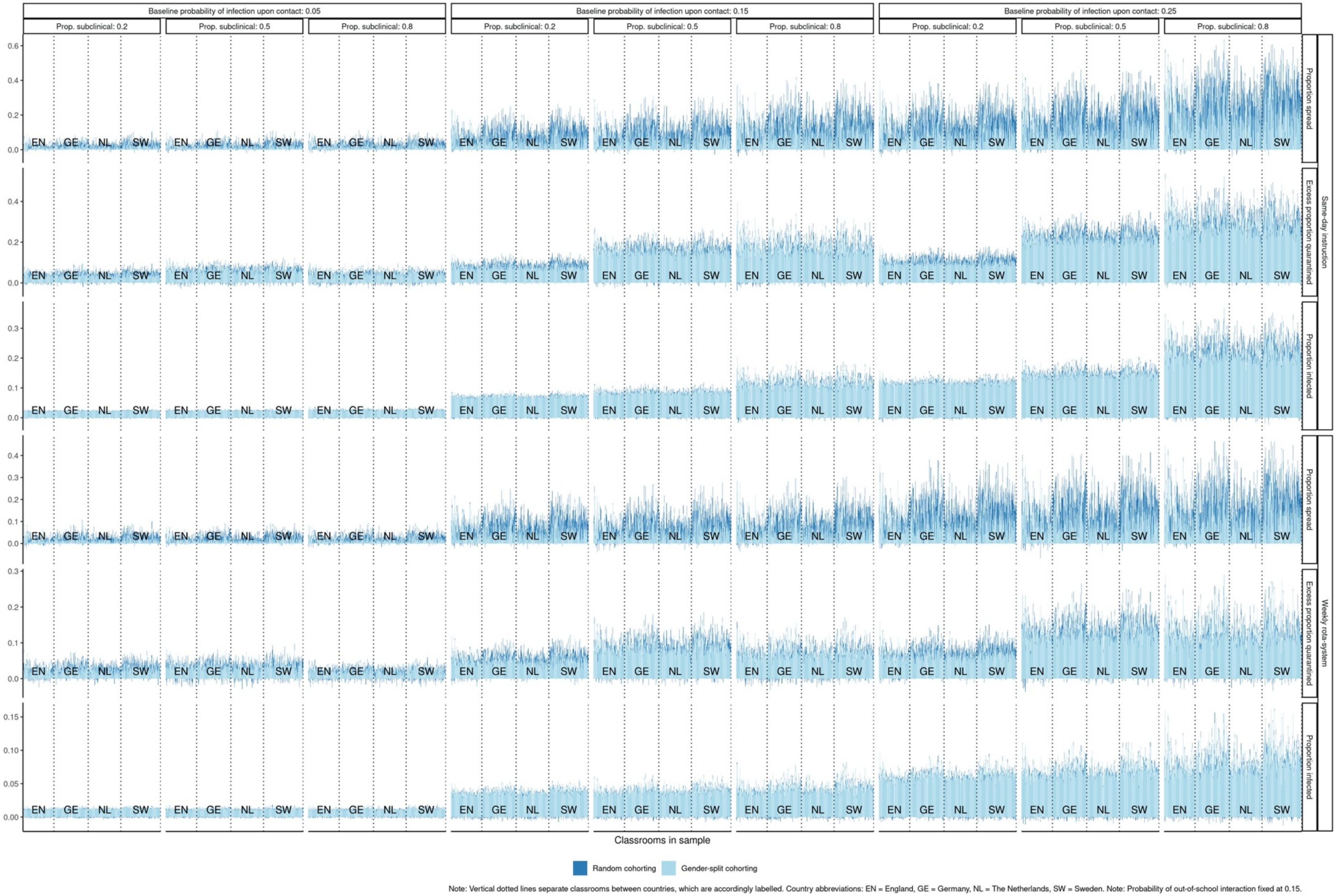


**Figure B4:** Random and gender-split cohorting strategies. Epidemiological outcomes for each classroom: Proportion of instances of infection spreading to the second cohort, proportion of students infected, and excess proportion of students quarantined. Cumulative proportions.

### C. Epidemiological outcomes for lower infectiousness of subclinical relative to clinical infections (30%)

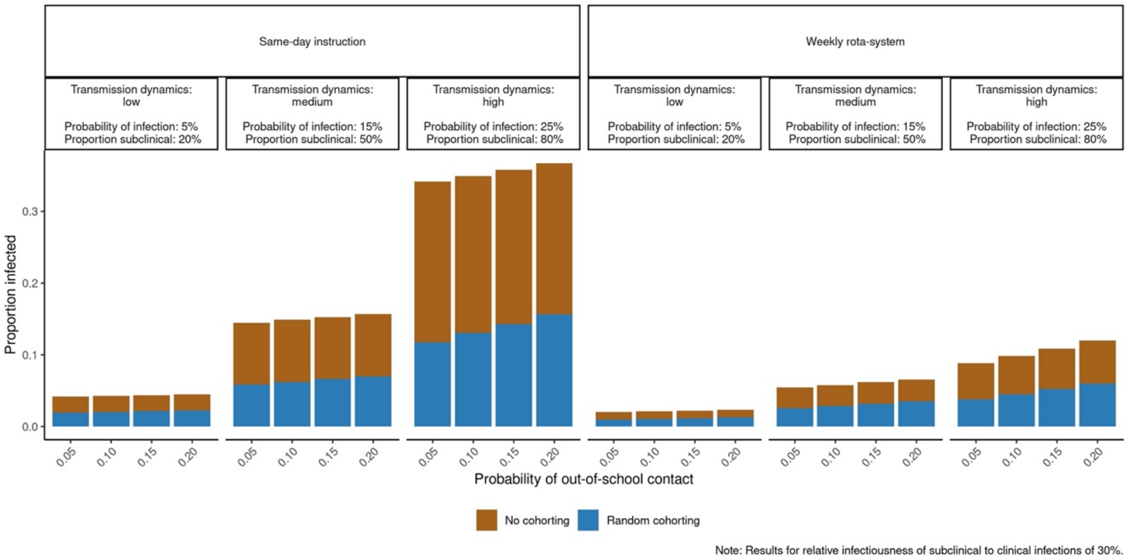


**Figure C1:** Lower infectiousness of subclinical relative to clinical infections (30%). Average cumulative proportion of infected classroom members in case of random cohorting (blue) and no cohorting (brown + blue)

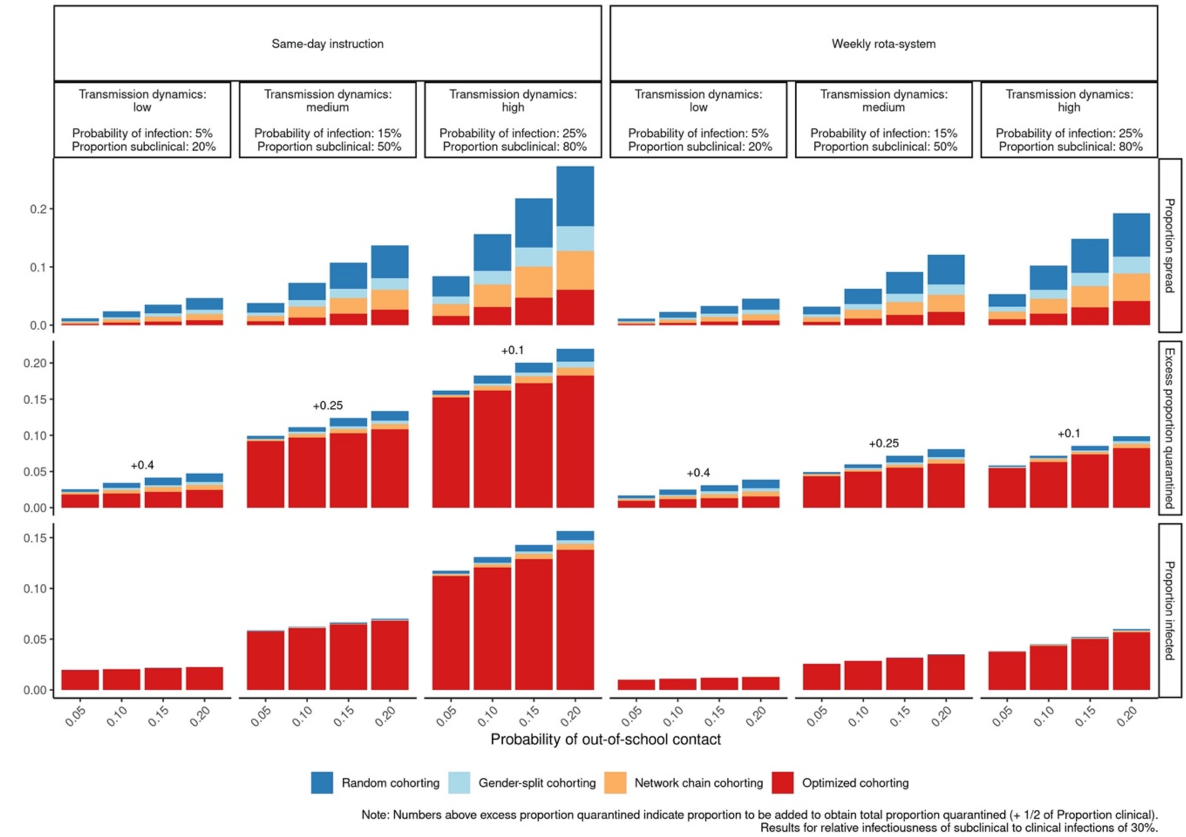


**Figure C2:** Lower infectiousness of subclinical relative to clinical infections (30%). Epidemiological outcomes of different cohorting strategies: Proportion of instances of infection spreading to the second cohort, proportion of students infected, and excess proportion of students quarantined. Cumulative proportions.

### D. Epidemiological outcomes for different proportions of in-classroom high-risk interactions (50% in Fig. D1 and 12.5% in Fig. D2)

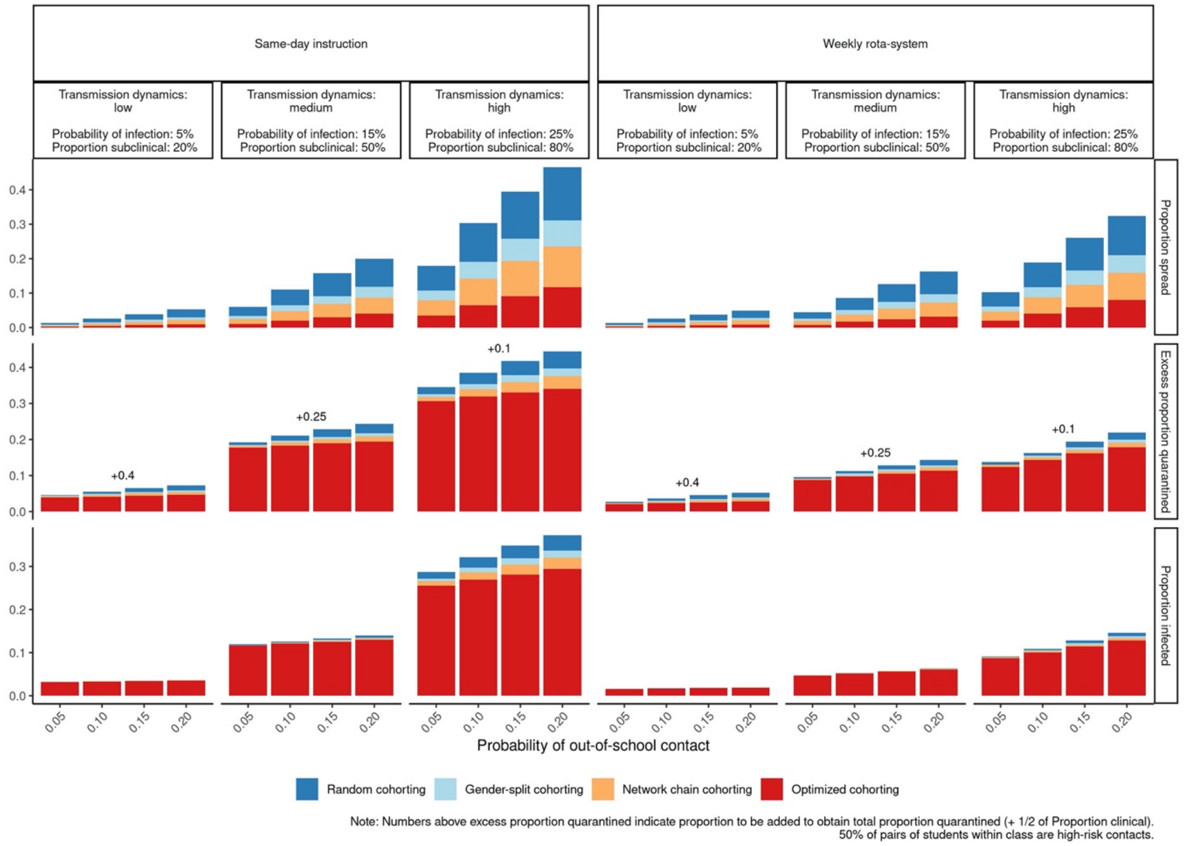


**Figure D1:** 50% High-risk interactions in classrooms. Epidemiological outcomes of different cohorting strategies: Proportion of instances of infection spreading to the second cohort, proportion of students infected, and excess proportion of students quarantined. Cumulative proportions.

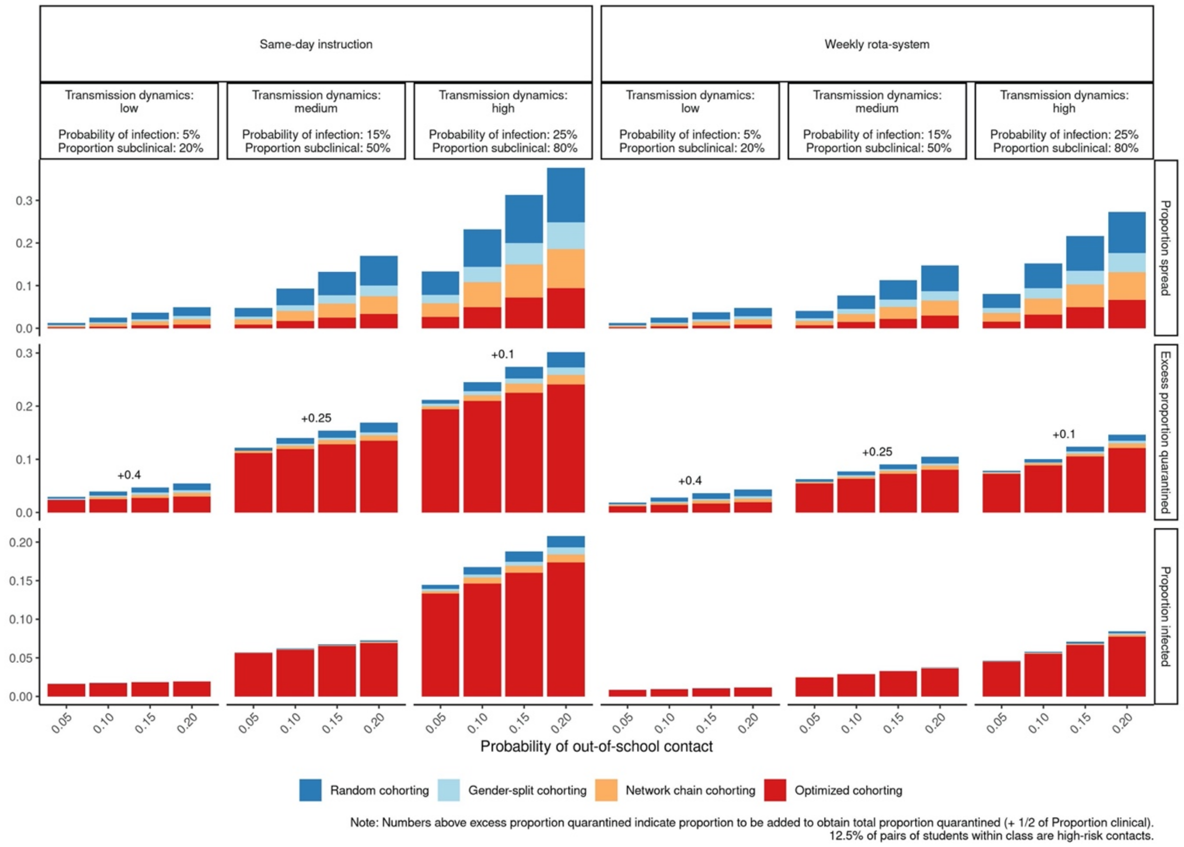


**Figure D2:** 12.5% High-risk interactions in classrooms. Epidemiological outcomes of different cohorting strategies: Proportion of instances of infection spreading to the second cohort, proportion of students infected, and excess proportion of students quarantined. Cumulative proportions.
